# Effects of Administration of Recombinant Human Lecithin-Cholesterol Acyltransferase on Lipoprotein Metabolism in Humans

**DOI:** 10.1101/2023.06.20.23291644

**Authors:** Gissette Reyes-Soffer, Anastasiya Matveyenko, James Lignos, Nelsa Matienzo, Leinys S Santos Baez, Antonio Hernandez-One, Lau Yung, Renu Nandakumar, Sasha A. Singh, Rajasekhar Ramakrishnan, Masanori Aikawa, Richard George, Henry N. Ginsberg

## Abstract

Lecithin cholesterol acyl transferase (LCAT) catalyzes the conversion of unesterified, or free cholesterol (FC), to cholesteryl ester (CE), which moves from the surface of HDL into the neutral lipid core. As this iterative process continues, nascent lipid-poor HDL is converted to a series of larger, spherical cholesterol ester enriched HDL particles that can be cleared by the liver in a process that has been termed reverse cholesterol transport (RCT). We conducted a randomized, placebo controlled, cross-over study in 5 volunteers with ASCVD, to examine the effects of an acute increase of recombinant human (rh) LCAT via intravenous administration on the in vivo metabolism of HDL apolipoprotein (APO)A1 and APOA2, and the APOB100-lipoproteins, very low density (VLDL), intermediate density (IDL), and low density (LDL) lipoproteins. As expected, rhLCAT treatment significantly increased HDL CE content. This change did not affect the fractional clearance or production rates of HDL-APOA1 and HDL-APOA2. The metabolism of APOB100-lipoproteins was likewise unaffected. Our results suggest that an acute increase in LCAT activity drives greater flux of CE through the RCT pathway without altering the clearance and production of the main HDL proteins and without affecting the metabolism of APOB100-lipoproteins. Long-term elevations of LCAT might, therefore, have beneficial effects on total body cholesterol balance and atherogenesis.

## Introduction

Lecithin cholesterol acyl transferase (LCAT) is a glycoprotein containing 416 amino acids with a molecular mass of approximately 67 kDa. It is secreted from the liver into the circulation where it associates primarily with high density lipoproteins (HDL) and, to a lesser degree, low density lipoproteins (LDL) ^1^. Glomset et. al. first reported esterification of free cholesterol in human plasma in 1962 ^2^, leading to the hypothesis that this activity might be critical for the flux of cholesterol from peripheral tissues to the liver, a process later named reverse cholesterol transport (RCT) ^3^.

LCAT catalyzes the conversion of unesterified, or free cholesterol (FC) to cholesteryl ester (CE) on HDL by the transacylation of fatty acid from the sn-2 position of phosphatidylcholine to the 3-hydroxyl group on the A-ring of cholesterol. Because of the increased hydrophobicity of CE compared to FC, LCAT-generated CE moves from the surface of HDL particles into the neutral lipid core ^4^. The movement of FC from the surface of a nascent, discoidal pre-beta HDL, whose other components are phospholipid and apolipoprotein AI (APOA1), into the core, allows additional cellular FC to move onto the surface where it is a substrate for LCAT. As this iterative process continues, pre-beta HDL is converted to a series of larger, spherical alpha-HDL particles. CE carried in the core of HDL particles can be cleared by hepatic receptors, such as SR-B1 or transferred to apolipoprotein B (APOB)-lipoproteins by cholesterol ester transfer protein (CETP)^5, 6^.

Preclinical animal models have been utilized to investigate the effects of LCAT on lipoprotein metabolism and atherosclerosis. Initial studies in wild-type mice demonstrated that transgenic expression of human LCAT substantially raised HDL-cholesterol (HDL-C), but LCAT had no effect on, or increased, atherosclerosis in studies where the latter was specifically examined ^7, 8^. Importantly, atherosclerosis was reduced in LCAT-overexpressing mice that also had deletion of SR-B1 ^9^ or concomitant overexpression of CETP ^10^. Furthermore, in rabbits, which express CETP endogenously, transgenic overexpression of human LCAT protected against diet-induced atherosclerosis ^11^.

In a first-in-human study by Shamburek et. al., infusions of a recombinant human LCAT (ACP-501) into an individual with LCAT deficiency resulted in a rapid rise in HDL-C from very low baseline levels of <5 mg/dL to levels of 10-20 mg/dL by 8-12 hours post-infusion. LDL-C levels, which were also very low at 46 mg/dL, increased more slowly than HDL-C, peaking at levels between 60-80 mg/dL 2-3 days post-infusion ^12^. These same authors published a Phase I single-ascending dose trial of ACP-501 in individuals with stable coronary heart disease (CHD) and HDL-C levels of 30-50 mg/dL. Infusion of ACP-501 increased HDL-C levels 40-50% over baseline at 12-24 hours post-infusion ^13^. In the present study, we investigated the effects of infusing MEDI6012, a recently developed recombinant human LCAT, into individuals with stable atherosclerotic cardiovascular disease (ASCVD) and normal levels of endogenous LCAT. MEDI6012 (hereafter referred to as rhLCAT) has been shown to significantly increase plasma levels of HDL-C in single- and multiple-ascending dose studies ^14, 15^. Our goal was to characterize in detail how increases in circulating LCAT affect plasma lipid and lipoprotein levels, lipoprotein composition, and the metabolism of APOB100, APOA1, and APOA2.

## Methods

### Study Design

We performed a phase 2, single-center, placebo-controlled, double-blind, randomized crossover study to determine the effects of an acute increase in rhLCAT on plasma lipid and lipoprotein levels and composition, as well as the kinetics of APOB100 in very low-density lipoprotein(VLDL), intermediate density lipoprotein (IDL) and LDL, and both HDL-APOA1, and HDL-APOA2, in subjects with stable ASCVD, (NCT03773172). The study-**Supplemental Figure 1**. was powered to assess the changes in the fractional clearance rate (FCR) and the production rate (PR) of LDL-APOB100 post-treatment with rhLCAT versus placebo. The LDL-APOB100 FCR is defined as the fraction of the circulating pool of LDL-APOB100 cleared from the plasma each day. Secondary outcomes included the effects of rhLCAT on the FCRs and PRs of VLDL-APOB100- and IDL-APOB100, the FCRs and PRs of HDL-APOA1 and HDL-APOA2, and HDL and VLDL, IDL, LDL composition and size. The study was conducted at the Columbia University Irving Medical Center (CUIMC).

### Study Cohort

There were 10 subjects screened. The study was approved by the Columbia University Irving Medical Center (CUIMC) Institutional Review Board and registered at clinical trials.gov (NCT03773172). All subjects agreed and signed the informed consent form. Once consented, subjects were assessed for eligibility by completing a screening visit, which included a medical history, physical exam, and fasting blood sampling for lipid levels and for safety studies. Eligible subjects were enrolled within 28±4 days. Subjects completed 24-hour food recalls to assess any extreme diet habits. Seven subjects were enrolled and randomized, and five subjects completed both study periods. Safety monitoring was performed 28±4days after both the first and second study phases. Two subjects did not complete the study: one withdrew consent after completing phase 1, the other completed phase 1, but could not complete phase 2 due to site closure at the start of the Covid19 pandemic. Once enrolled and randomized, subjects were instructed in an American Heart Association (AHA) heart healthy diet; we used frequent follow-up telephone calls to obtain diet histories and to reinforce diet compliance over the duration of the study.

### Study Treatment Protocol

We administered rhLCAT or placebo as intravenous bolus injections: a loading dose of 300 mg recombinant protein or placebo followed 48 hours later by a 150 mg dose of recombinant protein or placebo. The doses and timing of injections were based on a Phase 2 study of rhLCAT that demonstrated steady-state levels of rhLCAT over at least 36 hours ^15^. The placebo and rhLCAT was prepared for injection by the research pharmacy at CUIMC and the pharmacy staff held the randomization code.

### Stable Isotope Administration and Metabolic Studies

Studies were conducted in the outpatient and inpatient research units of the Irving Institute for Clinical and Translational Research (IICTR) at CUIMC. Briefly, in the morning of Day 0 of each study period, blood samples were obtained in the outpatient research unit after a 12-hour overnight fast. Then the first dose of either active drug or placebo was administered by intravenous bolus at approximately 8am. Subjects were observed during the next 2 hours for physical signs of acute placebo or rhLCAT-induced adverse reactions and then discharged. They returned to the IICTR at approximately 12pm the next day (Day 1-post first dose) and were admitted to the inpatient research unit for 2 nights. At the time of admission, safety bloods were drawn and immediately sent to the hospital laboratory for stat processing to ensure that no adverse effects had occurred after the first dose of the placebo or rhLCAT. After admission, all meals were provided by the staff of the IICTR Nutrition Research Unit. On Day 1, the subjects were made NPO at 8pm. At 1am (Day 2) liquid feedings (every 2 hours; 16 feedings) were begun as part of a protocol that achieves a nutritional steady state before and post-administration of stable isotopes ^16^. The composition of this liquid formula is 57% carbohydrate, 18% fat, and 25% protein. Subjects received the 2^nd^ dose of placebo or rhLCAT exactly 48 hours after Day 0 - first dose). One hour after the second dose (Day 2, approximately 9am), blood samples were obtained (0 hour) for pre-stable-isotope measurement of various lipids and lipoproteins. Immediately after this blood draw, stable isotopes were administered to subjects for the study of lipid and lipoprotein kinetics. Intravenous boluses of ^2^H_3_-L-leucine (10 μmol/kg/body weight (BW)), Ring-^13^C_6_-L-phenylalanine (29.4 μmol/kg BW), and ^2^H_5_-glycerol (100 μmol/kg BW) were administered over a 10-minute period, followed by a constant intravenous infusion of ^2^H_3_-L-leucine (10 μmol/kg BW/hour) over 15 hours. Blood samples were collected at 0 (pre-bolus), 20, and 40 minutes, and at 1, 2, 4, 6, 8, 10, 12, 14, 15, 15.5, 16, 18, 21, and 24 hours. Subjects were discharged on Day 3 after eating a heart healthy breakfast. Subjects repeated this same protocol with the alternate treatment after a 6–8-week washout period. During the washout period, subjects were monitored and re–instructed in the guidelines of an AHA diet. Two nutritional assessments via 24-hour recalls ensured that all subjects maintained a constant healthy diet during the washout period between the first and second studies. An extra fasting blood sample on Day 0 of the second study period was obtained for stat lipid measurements to ensure that the subject’s plasma lipid levels were the same as they were at the start of the first study. A visual representation of the investigative protocol is depicted in **Supplementary Figure 1**.

### Laboratory Methods

Plasma and lipoprotein lipid levels were measured in the IICTR Biomarkers Laboratory with an automated analyzer using Roche Reagents. Levels of apolipoproteins were determined by validated ELISAs with the following antibodies: APOB100 (Allercheck), APOA1, APOA2, APOC3, and APOE (Abcam). Lipoproteins (VLDL, IDL, LDL, HDL) were isolated from the 17 timed-samples via sequential ultracentrifugation as previously described ^17^. The concentrations of triglycerides (TG) and cholesterol, as well as APOB100 in VLDL, IDL, and LDL, and APOA1 and APOA2 in HDL were determined by the methods described above.

The FC and CE content of HDL were determined using the Amplex Red Cholesterol Assay Kit (Invitrogen) which enables measurement of total cholesterol and FC, with the difference being CE. Plasma lathosterol, campesterol, and sitosterol, as markers of cholesterol synthesis and absorption, were measured as previously described ^18^. The size and particle number of lipoprotein fractions in plasma were determined by ion mobility analysis ^19^.

We performed fast protein liquid chromatography (FPLC) to determine the distribution of cholesterol and APOA1 across the lipoprotein spectrum in fasting plasma obtained from the 0-hour time-point (obtained one-hour after administration of the second dose of placebo or rhLCAT and just prior to injections of the stable isotope tracers). FPLC was performed by loading 200 microliters of plasma into an ÄKTA FPLC system (GE Healthcare) using a Superose 6 10/300 GL column (catalog number 29021596 by Cytiva) at a flow of 0.3mL/min: Forty 0.5mL fractions were collected. Total cholesterol was measured in each fraction using a commercial kit (FUJIFILM Wako Diagnostics U.S.A.) adapted to 96 well plates. Western blots were performed using an anti-APOA1 antibody (Abcam).

Gas chromatography mass spectrometry (GC-MS) was used to measure enrichment of ^2^H_3_-L-leucine and Ring-^13^C_6_-L-phenylalanine in VLDL-, IDL-, and LDL-APOB100 (Agilent 6890 GC and a 5973 MS using negative chemical ionization); and ^2^H_5_-glycerol in VLDL-TG (using positive chemical ionization after derivatization of glycerol to triacetin) in each study period using previously published protocols ^20^. Liquid chromatography mass spectrometry (LC-MS) ^21^ was used to measure enrichment of ^2^H_3_-L-leucine in both LDL- and HDL-APOA1 and APOA2 in each study period (for detailed methods, see Supplementary Data). LC-MS was also used to obtain abundance of endogenous LCAT during placebo and endogenous LCAT plus rhLCAT during rhLCAT treatments. The proteome of isolated HDL particles was analyzed by LC-MS (see Supplementary Data).

### Stable Isotope Enrichment Data Modeling

We determinedAPOB100 turnover in VLDL, IDL, and LDL using the ^2^H_3_-L-leucine and Ring-^13^C_6_-L-phenylalanine enrichment data from the constant infusion and the bolus injection, respectively. TG turnover in VLDL was determined using ^2^H_5_-glycerol. The data were fitted by a multicompartmental model we have used in our prior work ^22, 23^ with a computer program, Poolfit, developed by us ^24^, which solves the differential equations in closed form and computes the fits and parameter sensitivities as sums of exponentials, yielding FCRs of APOB100 in VLDL, IDL, and LDL, and of VLDL-TG. ApoB100 and TG were required to have the same structure for VLDL and the same rate constants for the VLDL pools, but with different mass distributions. PRs were calculated by multiplying fractional clearance rates by the pool size of each lipoprotein-apolipoprotein and TG (measured concentrations x plasma volume, assuming 45 ml/kg for plasma volume). We calculated HDL- and LDL-APOA1 and APOA2 FCRs by fitting the leucine enrichment (obtained by LC-MS, **Supplementary Table 1**) in plasma LDL- and HDL-APOA1 and HDL-APOA2 with single-pool models, with the precursor enrichment set at the plateau level of VLDL-APOB leucine enrichment in that study. We reported FCRs as pools/day and the PRs of HDL AI and A2 mg/kg/day.

### Statistical analysis

Paired t-tests were used to compare variables at the end of the placebo period with those at the end of the active medication period. Significance for the pre-specified endpoints of LDL APOB100 FCR and LDL APOB100 PR was set at p<0.05. Significance for all other endpoints was set at p<0.01.

## Results

Five participants with stable ASCVD completed both study phases; their demographics and plasma lipids at screening are presented in **Table 1**. Mean LDL-C and HDL-C at screening were 46±15 and 35±11 mg/dL, respectively (all subjects were receiving statin treatment chronically). Mean lipid levels for all participants were similar at the start of each study period (Day 0, pre-placebo or rhLCAT for each study phase), **Supplementary Table 2**. We measured LCAT abundance by LC-MS in HDL isolated by ultracentrifugation at 3 time-points during the 24 hours of sampling for the kinetic studies in both the placebo and the rhLCAT study phases. There was a clear increase in LCAT levels in all subjects post-rhLCAT administration, **Figure 1**.

**Figure 1.**
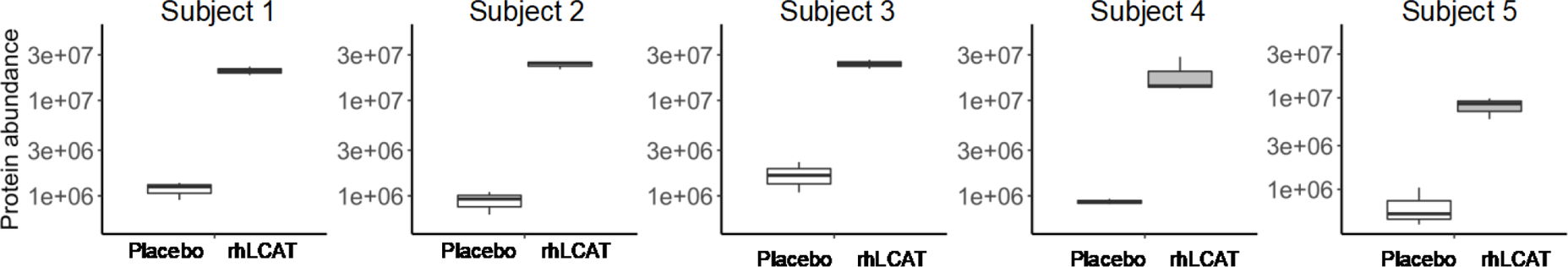
LCAT Abundances Increased in the rhLCAT Group Compared to the Placebo Treated Group. LCAT peptide abundance as determined by LC-MS-enabled proteomics. Abundances were measured in 3 timepoints per study subject during each study period (Placebo and rhLCAT). The rhLCAT data are placed second for ease of viewing; Subjects 1 and 5 received rhLCAT first in this randomized cross-over study.

**Table 1.** Study Population Characteristics. Ages are provided in 10 year ranges Blood samples for plasma lipids were obtained at screening visit, within 28 days of the start of the study. Chol-cholesterol; TG-triglycerides; LDL-low density lipoprotein; HDL-high density lipoprotein; All values in mg/dl

| Subject | 1 | 2 | 2 | 4 | 5 | Mean | SD |
| --- | --- | --- | --- | --- | --- | --- | --- |
| Sex | Male | Male | Female | Male | Male |  |  |
| Race | Caucasian | Caucasian | Hispanic | Hispanic | Caucasian |  |  |
| Age (10 year ranges) | 55-65 | 60-70 | 55-65 | 70-80 | 65-75 |  |  |
| Height (cm) | 172 | 182 | 152 | 160 | 185 | **170±14** | 14 |
| Weight (kg) | 74 | 83 | 75 | 62 | 84 | **75** | 9 |
| BMI (kg/m2) | 24.9 | 25.1 | 32.6 | 24.0 | 24.5 | **26.2** | 4 |
| Total Chol (mg/dL) | 95 | 116 | 104 | 76 | 108 | **100** | 15 |
| TG (mg/dL) | 90 | 83 | 103 | 113 | 90 | **96** | 12 |
| HDL-C (mg/dL) | 38 | 37 | 23 | 26 | 50 | **35** | 11 |
| LDL-C (mg/dL) | 39 | 62 | 61 | 28 | 40 | **46** | 15 |

As expected, treatment with rhLCAT increased total cholesterol (by 36.8±3.3 mg/dL; p<0.001) and HDL-C (by 34.9±10.3 mg/dL, p<0.001) levels compared to placebo, **Table 2**. Treatment with rhLCAT had no effects on plasma concentrations of LDL-C or TG when compared to placebo. Plasma APOB100 levels tended to be lower post-rhLCAT when compared to placebo (−15.5±9.0 mg/dL; p=0.04). APOA1 levels tended to be higher post-rhLCAT versus placebo compared to placebo (16.1±15.7 mg/dL; p=0.08). Concentrations of APOA2, APOE, and APOC3 did not change between periods, **Table 2**.

**Table 2.** Plasma Lipid and Apolipoprotein Levels during the Kinetic Studies on Placebo and rhLCAT Treatments. Chol-cholesterol; LDL-low density lipoprotein; HDL-high density lipoprotein; APO-apolipoprotein. Values are all mg/dl except for APOC3 and APOpoE, which are µg/dl. Values are the means of 3 time-points obtained during the 24 hour sampling period of the kinetic studies.

| **Plasma Measurements** | **Placebo Mean** | **Placebo SD** | **rhLCAT Mean** | **rhLCAT SD** | **Difference Mean** | **Difference SD** | **p-value** |
| --- | --- | --- | --- | --- | --- | --- | --- |
| **Total Chol** | 99.6 | 15.0 | 136.4 | 15.0 | 36.8 | 3.3 | < 0.001 |
| **Triglyceride** | 95.8 | 11.8 | 83.2 | 25.3 | -12.6 | 16.6 | 0.17 |
| **HDL Chol** | 34.7 | 10.8 | 69.6 | 20.0 | 34.9 | 10.3 | < 0.001 |
| **LDL Chol** | 45.8 | 14.9 | 50.2 | 17.4 | 4.4 | 7.1 | 0.24 |
| **APOB** | 67.4 | 15.3 | 51.9 | 6.5 | -15.5 | 9.0 | 0.04 |
| **APOA1** | 106.2 | 22.4 | 122.2 | 35.8 | 16.1 | 15.7 | 0.08 |
| **APOA2** | 31.1 | 5.7 | 31.8 | 6.8 | 0.7 | 5.8 | 0.81 |
| **APOC3** | 114.0 | 23.9 | 136.2 | 35.3 | 22.2 | 26.0 | 0.13 |
| **APOE** | 101.1 | 19.8 | 114.2 | 35.0 | 13.0 | 16.6 | 0.15 |

There were no significant differences in the concentrations of cholesterol and TG within isolated VLDL or IDL particles post-rhLCAT treatment when compared to placebo. LDL-TG had a trend toward a lower level after rhLCAT, but no changes in LDL cholesterol content were observed, **Supplementary Table 3**. The modest changes observed in the levels of plasma lipids and isolated APOB100-lipoproteins post-rhLCAT treatment were concordant with the results of the *in vivo* stable isotope kinetic studies of APOB100 metabolism, which demonstrated no effect of rhLCAT treatment on VLDL- or IDL-APOB100 metabolism. There was a trend for a lower level of LDL-APOB100 concentration during rhLCAT administration (p=0.05), but no clear differences for either of the two study primary endpoints, LDL-APOB100 FCR and LDL-APOB100 PR, during rhLCAT administration compared to placebo. Finally, rhLCAT administration did not alter VLDL-TG level, FCR, or PR, **Table 3**. Overall, these results indicated that significantly increasing LCAT mass over a three-day period did not result in significant alterations in the metabolism of APOB100-lipoproteins.

**Table 3.** VLDL-, IDL-, and LDL-APO B100 Fractional Clearance Rates and Production Rates on Placebo or rhLCAT Treatments. VLDL-very low density lipoprotein; IDL-intermediate density lipoprotein; LDL-low density lipoprotein; TG-triglycerides; APOB100 and TG in mg/dl; FCR – fractional clearance rate in pools/day; PR - production rate in mg/kg/day. APOB100 levels are the means of 3 time-points obtained during the 24 hrs of the kinetic studies.

| **Lipoprotein Fractions** | **Placebo Mean** | **Placebo SD** | **rhLCAT Mean** | **rh LCAT SD** | **Difference Mean** | **Difference SD** | **p-value** |
| --- | --- | --- | --- | --- | --- | --- | --- |
| **VLDL-APOB100 level** | 7.4 | 5.0 | 4.3 | 4.1 | -3.1 | 4.8 | 0.23 |
| **VLDL-APOB FCR** | 10.0 | 5.5 | 14.0 | 6.0 | 4.0 | 9.5 | 0.40 |
| **VLDL-APOB PR** | 29.6 | 20.3 | 19.9 | 11.4 | -9.7 | 23.5 | 0.41 |
| **IDL-APOB level** | 0.7 | 0.4 | 1.7 | 1.7 | 1.0 | 1.5 | 0.22 |
| **IDL-APOB FCR** | 5.2 | 2.6 | 6.7 | 2.1 | 1.5 | 3.7 | 0.42 |
| **IDL-APOB PR** | 1.8 | 1.8 | 4.8 | 5.2 | 3.0 | 3.67 | 0.14 |
| **LDL-APOB level** | 59.3 | 15.5 | 45.9 | 5.3 | -13.4 | 11.1 | 0.05 |
| **LDL-APOB FCR** | 0.5 | 0.3 | 0.8 | 0.2 | 0.3 | 0.3 | 0.13 |
| **LDL-APOB PR** | 14.2 | 11.3 | 15.7 | 3.2 | 1.5 | 11.0 | 0.78 |
| **VLDL-TG level** | 76.0 | 25.9 | 89.8 | 29.9 | 13.8 | 27.2 | 0.32 |
| **VLDL-TG FCR** | 10.0 | 5.5 | 14.0 | 6.0 | 4.0 | 9.5 | 0.40 |
| **VLDL-TG PR** | 371.6 | 283.8 | 536.8 | 224.9 | 165.2 | 389.7 | 0.40 |

In contrast to the minimal effects of rhLCAT treatment on markers of the composition and metabolism of APOB100-lipoproteins, HDL composition and metabolism were altered significantly. First, consistent with the data in Table 2, HDL-C isolated by ultracentrifugation doubled across the 5 subjects, increasing from a mean of 25.2±7.7 mg/dl to 53.0±16.6 mg/dL (p=0.003) during rhLCAT treatment compared to placebo**, Supplementary Table 3**. Importantly, this increase was due completely to a rise in HDL-CE content, **Figure 2**. Increased HDL-CE might have been associated with an increase in the number of HDL particles, an increase in HDL particle size, or both. As shown in **Table 2**, plasma levels of APOA1 tended to be higher during rhLCAT treatment whereas levels of APOA2 were not affected. Importantly, the marked increase in the CE content of HDL during rhLCAT administration was not associated with significant effects on HDL-APOA1 concentration, PR, or FCR, **Table 4**. The same was true for HDL-APOA2: there were no significant differences in the metabolism of HDL-APOA2 between rhLCAT and placebo treatment periods. Together, the steady state and stable isotope kinetic data suggest no significant rhLCAT-induced differences in the number of HDL particles or their metabolism, consistent with an isolated rhLCAT-mediated increase in the size of HDL.

**Figure 2.**
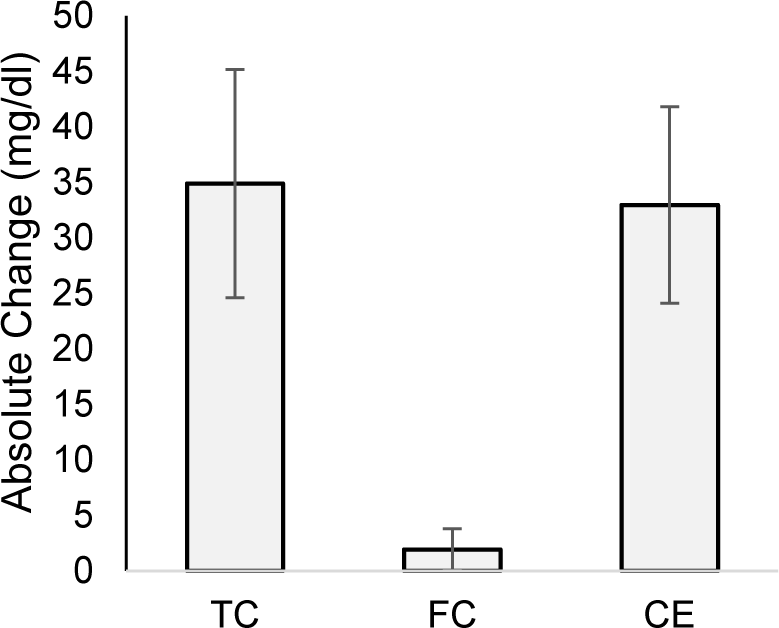
rhLCAT Treatment Increased Total HDL Cholesterol by Increasing Cholesterol Eester (CE). FC and CE content of HDL were determined using the Amplex Red Cholesterol Assay Kit which enables measurement of total cholesterol and FC, with the difference being CE. Data shown as the absolute differences (mg/dl) in TC, FC and CE between rhLCAT treatment and placebo.

**Table 4:**
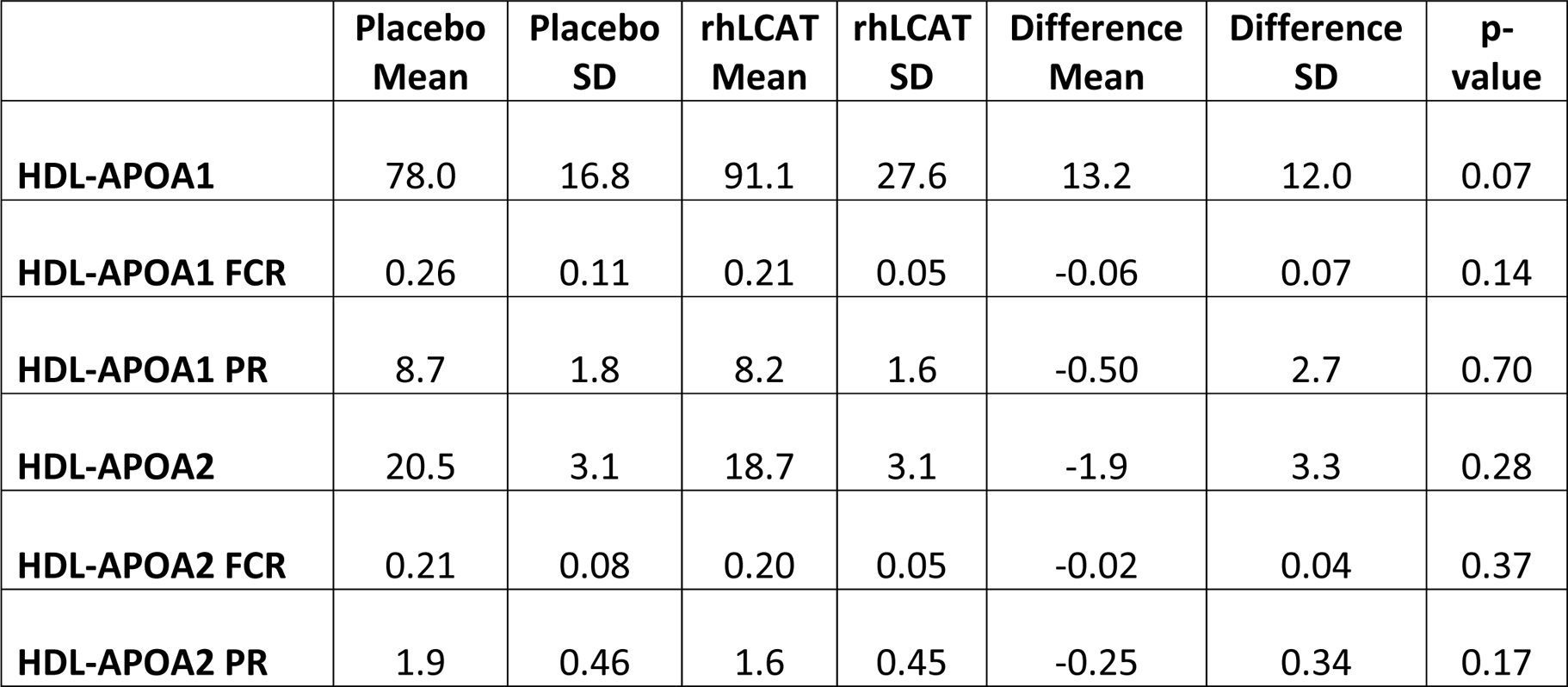
HDL-APOA1 and HDL-APOA2 Fractional Clearance Rates and Production Rates on Placebo or rhLCAT Treatments. HDL-high density lipoprotein; FCR – fractional clearance rate in pools/day; PR - production rate in mg/kg/day, APO-apolipoprotein. APOA1 and APOA2 levels are the means of 3 time-points obtained during the 24 hours of the kinetic studies.

To examine in more depth any change in HDL particle size, we used two independent approaches: ion mobility and chromatographic separation. Ion mobility, which measures both lipoprotein size and number directly, revealed increases in the number of large HDL2B particles concomitant with decreases in small HDL3_2a particles**, Supplementary Table 4**. Overall, there was a shift in the size of HDL lipoproteins toward larger, less dense particles, consistent with the increase in measured HDL-CE, **Figure 1**. Importantly, the combined concentration of HDL3-2a plus HDL2b particles were very similar, 17,053 and 16,783 nmol/L during the placebo and rhLCAT treatment periods, respectively, consistent with the absence of significant changes in APOA1 and APOA2 concentrations. Overall, therefore, rhLCAT treatment increased the CE content of HDL (and thus decreased its density) without altering the total number of HDL particles. There were also decreased numbers of mid-sized LDL particles (LDL IIab, LDL IIIab, and LDL IVa) with increases in the smallest (LDL IVb,c) and largest (LDL I) size particles. There was a modest reduction in total LDL concentration from 485 during placebo to 432 nmol/L during rhLCAT treatment, consistent with modest trends for reductions in plasma and LDL APOB100 levels described above. There were no changes in IDL or VLDL particle numbers.

Sizing of all circulating lipoproteins was determined using FPLC in a subgroup in whom there was available samples; a representative result is depicted in **Supplementary Figure 2**. FPLC analyses demonstrated an obvious shift of APOA1 and cholesterol from the HDL size-range into the LDL size-range in this subject during rhLCAT administration compared to placebo. These findings led us to examine the FCRs of APOA1 and APOA2 on lipoprotein particles that, because of CE-enrichment during administration of rhLCAT, were isolated by ultracentrifugation in the LDL density range. The results demonstrated that, like what was observed for the FCRs of the two proteins in HDL, there was no effect of rhLCAT on the FCRs of APOA1 and APOA2 isolated in the LDL density-range, **Supplementary Table 5**. Additionally, the FCRs for APOA1 and APOA2 isolated in the LDL density-range were not significantly different from the FCRs of each apoprotein isolated in the HDL density-range whether on placebo or rhLCAT (**Table 4**). Together, these analyses indicate that large, less dense CE-enriched APOA1/APOA2-lipoprotein particles, generated during administration of rhLCAT, had metabolic characteristics that were not different from typical HDL particles.

To investigate whether 3 days of increased LCAT activity achieved by administration of rhLCAT could increase RCT, we examined markers of whole-body cholesterol metabolism. First, we determined if there was suppression of endogenous cholesterol synthesis markers. We did not observe significant differences in levels of plasma lathosterol, a validated marker of cholesterol synthesis (p=0.9) between the two treatment periods. Additionally, there was no effect of rhLCAT administration on cholesterol absorption, assessed by measurement of plasma campesterol (p=0.8) and beta-sitosterol (p=0.7), **Supplementary Table 6**. These results indicate that a short-term increase in LCAT mass, which resulted in a doubling of HDL-CE mass, did not significantly affect total body cholesterol metabolism.

We assessed the effects of rhLCAT treatment on the proteome of ultracentrifugally isolated HDL and identified 71 proteins, **Supplementary Tables 7**). Proteins that were differentially abundant (p≤0.05; q≤0.10) between the placebo and rhLCAT treatment are presented as a heat map, **Figure 3A**. Including LCAT, 16 proteins were higher in abundance in the rhLCAT group, and 18 were higher placebo group. APOA1 and APOA2 tended to be lower in the rhLCAT group (p=0.18 and 0.09, respectively). These differentially abundant proteins represent distinct functions of HDL. For example, when all 71 HDL proteins were queried into the Gene Ontology database, the top-5 biological processes included ‘high-density lipoprotein particle remodeling”, “triglyceride-rich lipoprotein particle remodeling”, and “cholesterol transport”, processes linked to, for example, LCAT, PLTP, APOE, as well as “platelet degranulation” and “regulated exocytosis”, processes linked to, for example, serine protease inhibitors (SERPINs) and fibrinogens (FGA,FGB), **Figure 3B and Supplementary Table 8**. The division of the HDL proteome into two main functional groups, 1) lipid and cholesterol metabolism and 2) responses to vascular wounding and repair have been reported previously ^25^. In this study, four of the top-5 biological processes for proteins whose abundances were higher in the placebo group were related to vascular wounding and repair; however, all top-5 biological processes for the proteins whose abundances were higher in the rhLCAT group were related to lipid and cholesterol metabolism, **Figure 3B**.

**Figure 3.**
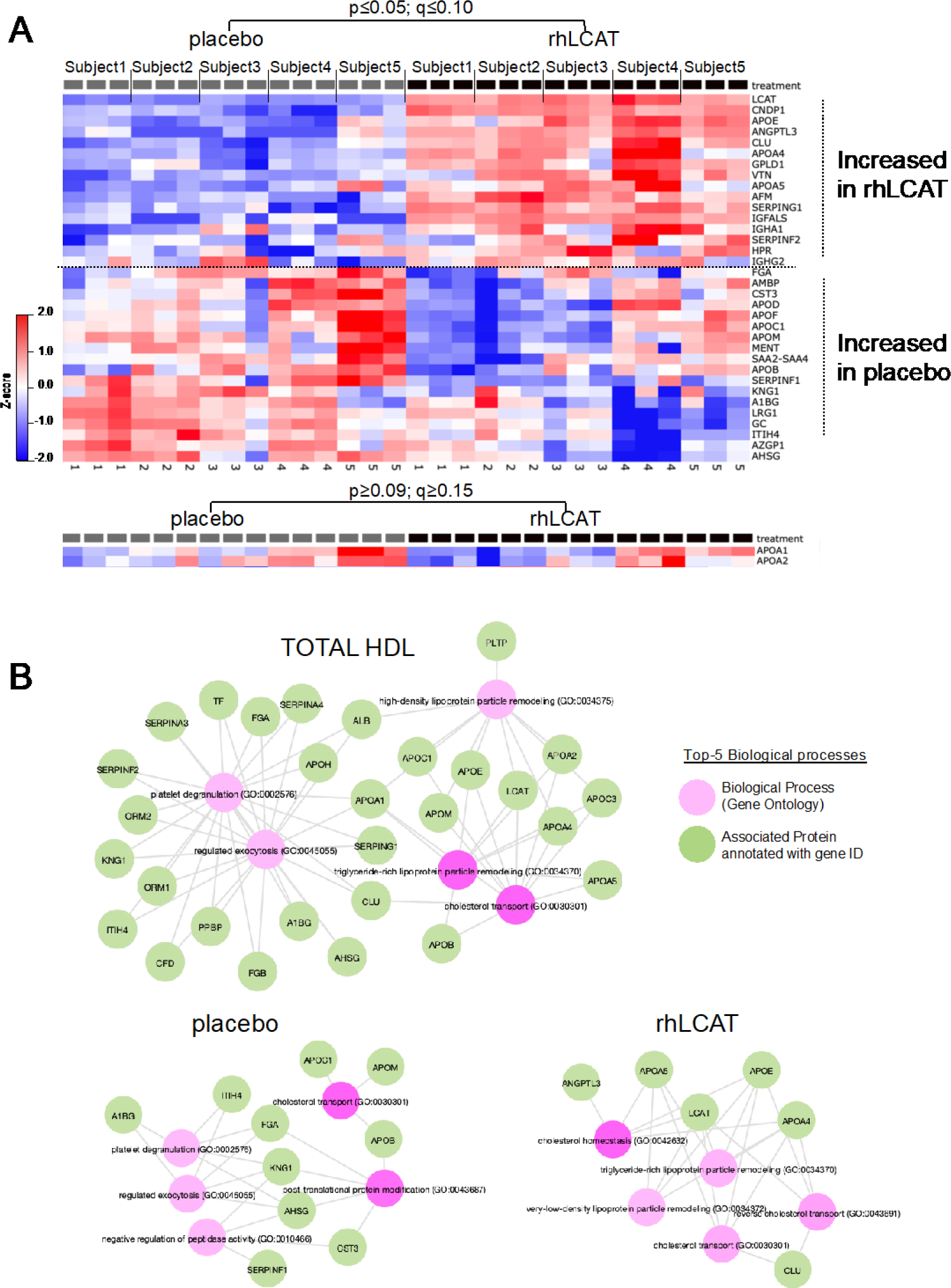
Effects of rhLCAT Treatment on the HDL Proteome. **A,** Heat map of HDL proteins that are differentially abundant in placebo versus rhLCAT treatment periods (p≤0.05; q≤0.10). APOA1 and APOA2 decreased slightly during rhLCAT treatment. The data shown for each subject is the mean of three samples obtained during the kinetic study. p was calculated using the student’s t-test of each treatment group’s mean, and q was calculated using the Benjamini-Hochberg correction. **B,** The top-5 Gene Ontology biological processes associated total HDL proteins, proteins with increased abundance in the placebo group, and proteins with increased abundance in the rhLCAT group. Networks were created with EnrichR https://maayanlab.cloud/Enrichr/.

## Discussion

The concept of RCT ^3–6^ as a potential athero-protective pathway for maintaining whole body cholesterol balance has made it the focus of numerous investigations over decades ^5^. Here, we provide the results of a first-in-human study of the effects of rhLCAT administration on the *in vivo* metabolism of both APOB100-lipoproteins and HDL-APOA1/APOA2-lipoproteins. In this study, we have addressed two important questions: 1) can these results inform us about the athero-protective potential of elevating plasma HDL-C levels by increasing LCAT activity?; 2) will comparison of the present results with those from similar stable isotope studies of the effects of CETP-inhibitors, a class of drugs that to date have had very modest or no anti-atherogenic activity ^26–28^, allow prediction of outcomes of trials with rhLCAT molecules?

To answer these questions, we will first review the effects of rhLCAT we observed on the metabolism of atherogenic APOB100 lipoproteins. Overall, the administration of rhLCAT to 5 subjects with stable ASCVD who all were receiving aggressive LDL-C lowering treatments had no significant effects on the levels of plasma LDL-C, or on the levels of cholesterol and TG in VLDL, IDL, and LDL isolated by ultracentrifugation. Levels of APOB100 in plasma and in LDL tended to be lower during treatment with rhLCAT compared to placebo (p=0.04 and p=0.05, respectively), but these did not reach significance as secondary endpoints. Importantly, neither of the pre-specified primary endpoints, LDL-APOB100 FCR or LDL-APOB100 PR, was different between the rhLCAT and placebo treatment periods.

These results differ from those seen for APOB100-lipoproteins in rabbits overexpressing human LCAT, where lower levels of non-HDL cholesterol were observed in association with increased FCRs of LDL ^29^; the basis for the difference is unclear. In the Phase 1 trial with ACP-501 ^13^, infusion of LCAT was associated with a very early modest fall in LDL-C followed by a modest rebound at 24 hours; these changes are consistent the absence of significant change in LDL-C across the 24-hour sampling period in our study. In the Phase 1 trial with ACP-501, there were no changes in overall mean TG or APOB100 during infusion of the protein, but no time-course data were provided. In addition, there were no effects of ACP-501 infusion on LDL particle number assessed by nuclear magnetic resonance. Our static concentrations of LDL-C, TG, APOB100, and ion mobility measures of LDL particle number following infusion of rhLCAT are consistent with those Phase I data. Importantly, two recent early phase trials with MEDI6012, the rhLCAT used in the present study, demonstrated modest increases in LDL-C and modest decreases in LDL-APOB100 associated with significant reductions in small LDL measured by NMR ^14, 15^. Our present study, in which similar, but non-significant trends for LDL-C and LDL-APOB100 concentrations were observed, adds the critically informative data that LDL-APOB FCR and PR were unaffected by rhLCAT treatment, despite clear evidence for LCAT-mediated increases in the generation of HDL-CE. In terms of increasing levels of plasma LCAT with the goal of reducing atherosclerosis risk, our results indicate that the metabolism of atherogenic APOB100-lipoproteins is not affected significantly by increases in LCAT activity. Of note, we previously demonstrated that healthy volunteers had significant reductions in LDL-C and LDL-APOB100 when treated with the CETP inhibitor anacetrapib, a drug that like rhLCAT, significantly increased plasma HDL-C concentrations ^16^. The reductions in LDL-C and APOB100 during treatment with anacetrapib were associated with significant increases in LDL-APOB FCR and this might explain the modest ASCVD benefit seen in the anacetrapib outcomes trial ^27^.

What can we learn from the effects of rhLCAT administration on the metabolism of HDL? As expected from both pre-clinical and limited clinical studies published to date, rhLCAT-mediated increases in LCAT levels were associated with changes in HDL levels, size, and composition. Thus, CE enrichment of HDL resulted in a concomitant shift in the profile of HDL from the smaller, denser HDL3_2a to the larger, less dense HDL2b subpopulation. In pre-clinical investigations of mice and rabbits, transgenic expression of human LCAT resulted in dramatic increases in HDL-C associated with CE-enrichment and increased HDL size ^29–32^. Administration of ACP-501 to an LCAT-deficient patient caused rapid decreases in small HDL subpopulations and the appearance of large, CE-rich HDL particles ^12^. Similar changes in HDL-C and HDL subpopulations were observed in the single-dose escalation, Phase I trial of ACP-501 in subjects with stable cardiovascular disease ^13^. In the two early phase trials with MEDI6012 referred to above ^14, 15^, increases in HDL-C, HDL-CE, APOA1, *ex vivo* measures of ABCA1-mediated cholesterol efflux from macrophages, and decreases in pre-beta HDL particles, were observed over several days following administration of rhLCAT.

In pre-clinical investigations of transgenic rabbits over-expressing human LCAT, changes in HDL composition and size were associated with a reduced plasma FCR of rabbit HDL-APOA1 compared to control rabbits ^29, 32^. Human LCAT transgenic mice had larger HDL that were associated with a significantly lower plasma FCR of HDL-[^3^H]CE than control mice; the LCAT transgenic mice also had less hepatic uptake of [^3^H]CE ^10^. A study of 5 patients with LCAT deficiency showed greater FCRs for both APOA1 and APOA2 compared to FCRs in normal controls ^33^. Tracer kinetic studies were not conducted in the trials of ACP-501 administration to one LCAT-deficient patient nor to the stable CHD subjects described above ^12, 13^. In the present study, both plasma and HDL-APOA1 concentrations trended toward increased levels during administration of rhLCAT compared to placebo, whereas no change in APOA2 was observed. Importantly, there were no significant changes in the FCR or PR of either HDL-APOA1 or HDL-APOA2 during infusion of rhLCAT compared to placebo.

Although the exact basis for the differences observed in pre-clinical studies and our clinical study is not completely understood, there are some differences in the studies that could explain these findings. First, unlike humans, mice lack CETP, and the importance of this protein in providing a pathway for RCT that would be in addition to the direct interaction of HDL with the liver was demonstrated by crossing CETP transgenic mice with LCAT transgenic mice. Compared to LCAT transgenic mice, the LCAT/CETP transgenic mice had lower levels of HDL-C and a greater FCR for HDL-CE as well as greater uptake of [^3^H]CE by the liver ^10^. Second, it must be noted that increased LCAT mass and activity were present from birth in the transgenic animals compared to an acute increase over 48 hours during our study protocol; lifetime increases in LCAT may have secondary effects to reduce the FCRs of HDL-APOA1 and HDL-APOA2. On the other hand, our protocol was of adequate duration to result in increases in HDL-C, HDL-CE, and HDL size that were like those seen in transgenic mice and rabbits.

Regarding the potential of rhLCAT treatment to reduce atherosclerosis risk, our *in vivo* metabolic studies indicate that the flux of larger, CE-enriched HDL particles, as reflected by the kinetics of HDL-APOA1 and HDL-APOA2, was normal. Additionally, proteomic analyses demonstrated that the increased CE content and size of HDL during rhLCAT treatment were associated with parallel increases in proteins related to cholesterol and lipoprotein metabolism, indicating that these were metabolically normal HDL. This would support a conclusion that RCT may be increased by chronic elevations of plasma LCAT, leading to reduced risk of ASCVD. Importantly, prior studies of the effects of two different CETP inhibitors that both altered HDL levels, size, and composition like the changes we observed with rhLCAT, differed from the latter in their effects on APOA1 metabolism. Our previous study demonstrated a reduced FCR of HDL-APOA1 but not HDL-APOA2 during CETP inhibition with anacetrapib ^20^. A study of the CETP inhibitor torcetrapib also demonstrated a significant reduction in the FCR of APOA1; APOA2 kinetics were not reported ^35^.

## STUDY LIMITATIONS

The two major limitations of this study, in terms of drawing conclusions, are the small size of the study cohort and the short duration of rhLCAT administration. Regarding the small cohort of only five subjects, the start of the COVID-19 pandemic essentially shut down clinical research, particularly studies requiring in-hospital stays. Additionally, during the extended pandemic period, the sponsor ended their rhLCAT program. A review of our results suggests that the greater LDL-APOB100 FCR and the lower LDL-APOB100 concentration during infusion of rhLCAT may have reached statistical significance (p<0.05 and p<0.01, respectively) with a study cohort 2-3 times larger. Further, we agree that a higher LDL-APOB100 FCR together with a lower LDL-APOB100 concentration during rhLCAT would have been an intriguing outcome. Regarding the short duration of rhLCAT administration, the half-life of the recombinant protein required two infusions 48 hours apart to obtain a steady state long enough for us to conduct the 24-hour stable isotope kinetic protocol. If we wanted to have a longer treatment period prior to the start of the kinetic study, the subjects would have had to come to the IICTR research center every 48 hours for an extended time-period. In defense of the short duration of treatment, the expected effects of rhLCAT on HDL CE content, size, and density were all observed by 24 hours. Additionally, HDL-C, TG, APOB100, APOA1, and APOA2 levels were stable during the 24-hour kinetic study.

## CONCLUSION

In conclusion, this first-in-human study of the effects of rhLCAT on the metabolism of APOB100, APOA1, and APOA2, are consistent with a long-standing model for the key role of LCAT in the esterification of FC on small, disc-shaped HDLs allowing those nascent HDLs to become larger, spherical, CE-enriched particles that are required for effective RCT ^6^. Importantly, our detailed tracer kinetic data indicate that the increased transport of LCAT-derived CE in HDL does not affect the kinetics of either of the key HDL proteins, APOA1 and APOA2, nor the kinetics of the APOB100 lipoproteins, VLDL, IDL, and LDL. In contrast to the failure of torcetrapib. and the very modest effect of anacetrapib (both of which affected APOB100 and APOA1 metabolism ^16, 20, 34, 36^) on CVD events in high-risk cohorts ^26, 27^, our data allow for continued optimism that other pharmacologic or genetic manipulations of RCT, including increased LCAT activity, will demonstrate benefit ^36^. Specifically, increased LCAT activity, by accelerating the conversion of FC to CE at a very early stage of the RCT pathway without affecting downstream metabolism of CE-enriched HDL, should result in a net increase in RCT. In contrast, inhibiting CETP-mediated transfer of CE from mature HDL to APOB-lipoproteins may, as some have suggested, reduce the overall efficiency of RCT. Our optimism must be guarded, however, in view of the recent report of the failure of short–term treatment of acute coronary syndrome patients with MEDI6012, the rhLCAT used in the present study, to improve CVD outcomes ^37^. Future attempts to increase LCAT activity should focus on approaches that allow for chronic administration of agents either orally or by subcutaneous injection.

## Data Availability

All data produced in the present work are contained in the manuscript.

## Acknowledgements

Laboratory measurements of lipoprotein size were performed in the laboratory of Dr. Ronald Krauss; measurements of sterols were performed in the laboratory of Dr. Alice Lichtenstein. The authors wish to acknowledge the outstanding work by the nursing staff and members of the Nutrition Research Unit of the ICCTR.

## Supplementary Data

### Methods

#### Lipoprotein proteolysis

Proteolysis was automated using the PreON (PreOmics GmbH, Germany) with the iST (96x) columns (PreOmics). For HDL, 40 µl of the iST LYSE buffer was added to 10 µl sample, corresponding to 20 ug protein input and subsequent proteolysis steps (including a 70°C lysis for 12 minutes and a 2-hour digestion) were done on the PreON (12-24 samples a day). For LDL, 25 µl of the iST 2XLYSE buffer was added to 25 µl sample. Lyophilized peptides were re-suspended in 5% acetonitrile and 0.5% formic acid dissolved in MS-grade water.

#### Data-dependent acquisition (DDA) for lipoprotein proteome profiling

Mass spectra were acquired on the Orbitrap Lumos coupled to an Easy-nLC1000 HPLC pump (Thermo Fisher Scientific). The peptides (n=3 time points per subject per treatment group) were diluted 10-fold using sample loading buffer and 2 ul injections separated using a dual column set-up: an Acclaim PepMap RSLC C18 trap column, 75 µm X 20 mm; and a heated EASY-Spray column (45°C), 75 µm X 250 mm (Thermo Fisher Scientific). The gradient flow rate was 300 nl/min from 8 to 25% solvent B (0.5% formic acid in acetonitrile) for 50 minutes, 25 to 30% solvent B for 10 minutes, and then 30 to 95% solvent B for 5 minutes; followed by an additional 10 minutes of 95 to 5% solvent B jigsaw wash. Solvent A was 0.5% formic acid in mass spectrometry-grade water. The mass spectrometer was set to 120 K resolution, and the top N precursor ions in a 3 second cycle time (within a scan range of 375-1500 m/z) were subjected to higher energy dissociation (HCD, collision energy 30%) for peptide sequencing using a 30 K resolution setting. The parallelization feature was enabled (automatic gain control/AGC target, 1.0e5; maximum injection time, 54 ms).

#### Targeted mass spectrometry (tMS2) for ^2^H_3_-L-leucine tracer enrichment analysis

tMS2 was also performed on the Orbitrap Lumos with 2 ul injections of 10-fold diluted peptide stocks. Leucine-containing peptides were identified in the DDA analysis. The gradient flow rate was 300 nl/min from 8 to 25 % solvent B for 30 minutes, 25 to 95 % Solvent B for 2 minutes, followed by an additional 5 minutes of 95 % solvent B. tMS2 on 3 APOA1, 2 APOA2 and 3 APOB leucine-containing peptides was performed using the ‘targeted MS2 scan’ module in scheduled mode. The isolation window was set to 4 Da around the average of the M0 and 2HM3 precursor *m/z* values ^1^, **Supplemental Table 1**. Dissociation was set to 30% HCD collision energy, and the MS2 scan range (*m/z* 150-1000) was set to 240 K resolution (AGC target 2.0e5; maximum injection time, 502 ms).

#### Spectral processing

A total of 30 DDA datasets (n=5 subjects per placebo and rhLCAT treatment group X 3 timepoint replicates each) were queried against the Human UniProt database (downloaded August 01, 2020; 96,816 entries) using the HT-SEQUEST search algorithm, via the Proteome Discoverer (PD) Package (version 2.2, Thermo Fisher Scientific). The search criteria were as followings: a 10 ppm tolerance window in the MS1 search space, and a 0.02 Da fragment tolerance window for HCD data. Methionine oxidation was set as a variable modification, and cysteine carbamidomethylation was set as a static modification. The peptide false discovery rate (FDR) of 1% was calculated using Percolator provided by PD2.2. In order to quantify peptide precursors detected in the MS1 but not sequenced from DDA dataset to dataset, we enabled the ‘Feature Mapper’ node. Chromatographic alignment was done with a maximum retention time (RT) shift of 10 minutes and a mass tolerance of 10 ppm. Feature linking and mapping settings were, RT tolerance minimum of 0 minutes, mass tolerance of 10 ppm and signal-to-noise minimum of five. Precursor peptide abundances were based on their chromatographic intensities and total peptide amount was used for normalization. Peptides assigned to a given protein group, and not present in any other protein group, were considered as unique. Consequently, each protein group is represented by a single master protein (PD2.2 Grouping feature). Proteins with 2 or more unique peptides were exported. For tracer enrichment analysis, APOA1, APOA2 and APOB leucine-containing peptides were curated in Skyline ^2^, to calculate the light (M0) and heavy (2HM3) precursor and fragment ion peaks. Fragment ion peak intensities were manually calculated using XCalibur (Thermo Fisher Scientific) ^3^.

#### Proteome profiling and statistical analysis

The HDL proteome was exported from PD2.2, and a median normalization of each sample’s proteome was done. ^4, 5^. A two-group means analysis (Placebo vs. rhLCAT) was done using the statistical software Qlucore Omics Explorer (version 3.7; Qlucore, Sweden). P-values were calculated using the student’s t-test and the false discovery rate value (q-values) was calculated using the Benjamini-Hochberg correction ^6^ (Qlucore).

#### Gene Ontology Networks

The EnrichR tool was used to generate the protein-biological processes (from Gene Ontology Database) networks ^7^ for HDL, placebo and rhLCAT groups (Supplemental Table 8)https://maayanlab.cloud/enrichr-kg.

**Supplementary Table 1.** Peptide and Fragmentations Monitored for tMS2-based enrichment analysis

| protein | label | peptide | rt start | rt end | isolation mz (4Da window) | precursor mass | fragment mass | fragment | lipoprotein |
| --- | --- | --- | --- | --- | --- | --- | --- | --- | --- |
| ApoA1 | Leu-D0 | LLDNWDSVTSTFSK | 25 | 35 | 807.651 | 806.8963 | 342.2023 | b3 | HDL, LDL |
| ApoA1 | Leu-D3 | LLDNWDSVTSTFSK | 25 | 35 | 807.651 | 808.4057 | 345.2212 | b3 | HDL, LDL |
| ApoA1 | Leu-D0 | LLDNWDSVTSTFSK | 25 | 35 | 807.651 | 806.8963 | 456.2453 | b4 | HDL, LDL |
| ApoA1 | Leu-D3 | LLDNWDSVTSTFSK | 25 | 35 | 807.651 | 808.4057 | 459.2641 | b4 | HDL, LDL |
| ApoA1 | Leu-D0 | LLDNWDSVTSTFSK | 25 | 35 | 807.651 | 808.4057 | 642.3246 | b5 | HDL, LDL |
| ApoA1 | Leu-D3 | LLDNWDSVTSTFSK | 25 | 35 | 807.651 | 808.7076 | 645.3434 | b5 | HDL, LDL |
| ApoA1 | Leu-D0 | THLAPYSDELR | 13 | 16 | 435.057 | 434.5543 | 619.3046 | y5 | HDL, LDL |
| ApoA1 | Leu-D3 | THLAPYSDELR | 13 | 16 | 435.057 | 436.5669 | 622.3234 | y5 | HDL, LDL |
| ApoA1 | Leu-D0 | THLAPYSDELR | 13 | 16 | 435.057 | 434.5543 | 532.2726 | y4 | HDL, LDL |
| ApoA1 | Leu-D3 | THLAPYSDELR | 13 | 16 | 435.057 | 436.5669 | 535.2914 | y4 | HDL, LDL |
| ApoA1 | Leu-D0 | THLAPYSDELR | 13 | 16 | 435.057 | 434.5543 | 417.2456 | y3 | HDL, LDL |
| ApoA1 | Leu-D3 | THLAPYSDELR | 13 | 16 | 435.057 | 436.5669 | 420.2644 | y3 | HDL, LDL |
| ApoA1 | Leu-D0 | THLAPYSDELR | 13 | 16 | 435.057 | 434.5543 | 423.2350 | b4 | HDL, LDL |
| ApoA1 | Leu-D3 | THLAPYSDELR | 13 | 16 | 435.057 | 436.5669 | 426.2539 | b4 | HDL, LDL |
| ApoA1 | Leu-D0 | THLAPYSDELR | 13 | 16 | 435.057 | 434.5543 | 520.2878 | b5 | HDL, LDL |
| ApoA1 | Leu-D3 | THLAPYSDELR | 13 | 16 | 435.057 | 436.5669 | 523.3066 | b5 | HDL, LDL |
| ApoA1 | Leu-D0 | AKPALEDLR | 8.5 | 11 | 338.701 | 338.1977 | 645.3566 | y5 | HDL, LDL |
| ApoA1 | Leu-D3 | AKPALEDLR | 8.5 | 11 | 338.701 | 339.2040 | 648.3754 | y5 | HDL, LDL |
| ApoA1 | Leu-D0 | AKPALEDLR | 8.5 | 11 | 338.701 | 338.1977 | 532.2726 | y4 | HDL, LDL |
| ApoA1 | Leu-D3 | AKPALEDLR | 8.5 | 11 | 338.701 | 339.2040 | 535.2914 | y4 | HDL, LDL |
| ApoA1 | Leu-D0 | AKPALEDLR | 8.5 | 11 | 338.701 | 338.1977 | 403.2300 | y3 | HDL, LDL |
| ApoA1 | Leu-D3 | AKPALEDLR | 8.5 | 11 | 338.701 | 339.2040 | 406.2488 | y3 | HDL, LDL |
| ApoA2 | Leu-D0 | EQLTPLIK | 16 | 22 | 472.042 | 471.2869 | 571.3814 | y5 | HDL, LDL |
| ApoA2 | Leu-D3 | EQLTPLIK | 16 | 22 | 472.042 | 474.3058 | 574.4002 | y5 | HDL, LDL |
| ApoA2 | Leu-D0 | EQLTPLIK | 16 | 22 | 472.042 | 471.2869 | 470.3337 | y4 | HDL, LDL |
| ApoA2 | Leu-D3 | EQLTPLIK | 16 | 22 | 472.042 | 474.3058 | 473.3525 | y4 | HDL, LDL |
| ApoA2 | Leu-D0 | EQLTPLIK | 16 | 22 | 472.042 | 471.2869 | 373.2809 | y3 | HDL, LDL |
| ApoA2 | Leu-D3 | EQLTPLIK | 16 | 22 | 472.042 | 474.3058 | 376.2998 | y3 | HDL, LDL |
| ApoA2 | Leu-D0 | EQLTPLIK | 16 | 22 | 472.042 | 471.2869 | 371.1925 | b3 | HDL, LDL |
| ApoA2 | Leu-D3 | EQLTPLIK | 16 | 22 | 472.042 | 474.3058 | 374.2113 | b3 | HDL, LDL |
| ApoA2 | Leu-D0 | SPELQAEAK | 9 | 10 | 487.5081715 | 486.753464 | 427.21873 | b4 | HDL, LDL |
| ApoA2 | Leu-D3 | SPELQAEAK | 9 | 10 | 487.5081715 | 488.262879 | 430.23756 | b4 | HDL, LDL |
| ApoA2 | Leu-D0 | SPELQAEAK | 9 | 10 | 487.5081715 | 486.753464 | 659.3723 | y6 | HDL, LDL |
| ApoA2 | Leu-D3 | SPELQAEAK | 9 | 10 | 487.5081715 | 488.262879 | 662.3911 | y6 | HDL, LDL |

**Supplementary Table 2.** Lipid Levels at Start of each Study Period. Chol-cholesterol; TG-triglycerides; LDL-low density lipoprotein; HDL-high density lipoprotein. All values in mg/dl. Lipids were measured on Day 0 of each study period.

| **Phase I** | **Total Chol** | **TG** | **HDL Chol** | **LDL Chol** |
| --- | --- | --- | --- | --- |
| 1 | 113.0 | 62.0 | 43.5 | 57.1 |
| 2 | 127.7 | 67.0 | 43.1 | 71.1 |
| 3 | 113.2 | 100.0 | 23.1 | 70.1 |
| 4 | 92.8 | 109.0 | 32.0 | 39.0 |
| 5 | 122.3 | 63.0 | 59.8 | 49.9 |
| Mean | **113.8** | **80.2** | **40.3** | **57.4** |
| SD | 13.3 | 22.5 | 13.8 | 13.6 |
| Phase II |  |  |  |  |
| 1 | 118.6 | 75.0 | 42.7 | 60.9 |
| 2 | 116.4 | 63.0 | 38.5 | 65.3 |
| 3 | 115.1 | 108.0 | 26.8 | 66.7 |
| 4 | 90.7 | 112.0 | 29.6 | 38.7 |
| 5 | 155.2 | 90.0 | 79.0 | 58.2 |
| Mean | **119.2** | **89.6** | **43.3** | **58.0** |
| SD | 23.1 | 21.0 | 21.0 | 11.3 |

**Supplementary Table 3.** Cholesterol and Triglyceride Content within Isolated Lipoprotein Fractions. VLDL-very low density lipoprotein; IDL-intermediate density lipoprotein; LDL-low density lipoprotein; HDL-high density lipoprotein; Chol-cholesterol. Values were the means of levels in 3 samples obtained during the 24 hour kinetic study. All values in mg/dl

|  | Placebo<br>Mean | Placebo<br>SD | rhLCAT<br>Rx<br>Mean | rhLCAT<br>Rx SD | Difference<br>Mean | Difference<br>SD | p-value |
| --- | --- | --- | --- | --- | --- | --- | --- |
| **VLDL Chol** | 9.8 | 5.1 | 8.0 | 4.7 | -1.8 | 6.0 | 0.53 |
| **VLDL Triglycerides** | 57.4 | 30.5 | 42.3 | 21.0 | -15.1 | 26.1 | 0.26 |
| **IDL Chol** | 2.2 | 1.3 | 3.1 | 2.6 | 0.9 | 2.2 | 0.40 |
| **IDL Triglycerides** | 5.9 | 3.8 | 5.4 | 4.1 | -0.4 | 1.4 | 0.53 |
| **LDL Chol** | 32.9 | 14.4 | 38.9 | 13.3 | 6.0 | 23.8 | 0.60 |
| **LDL Triglycerides** | 10.3 | 2.0 | 7.5 | 1.8 | -2.8 | 2.0 | 0.04 |
| **HDL Chol** | 25.2 | 7.7 | 53.0 | 16.6 | 27.8 | 9.6 | 0.003 |
| **HDL Triglycerides** | 8.3 | 0.9 | 7.4 | 1.6 | -0.9 | 2.0 | 0.37 |

**Supplementary Table 4.** Lipoprotein Particle Numbers During Placebo and rhLCAT treatments. Lipoprotein particle numbers were determined by ion-mobility in the laboratory of Dr. Ronald Krauss ^19^. All values in nmol/L.

| nmol/L | Placebo | SD | rhLCAT | SD | pvalue |
| --- | --- | --- | --- | --- | --- |
| **HDL 3_2a** | 12826.47 | 1758.38 | 9751.97 | 1049.32 | <0.01 |
| **HDL 2b** | 4227.02 | 989.42 | 7033.29 | 1492.34 | 0.01 |
| **LDL IVc** | 55.12 | 7.93 | 81.84 | 17.82 | 0.02 |
| **LDL IVb** | 52.39 | 6.73 | 57.31 | 13.70 | 0.49 |
| **LDL IVa** | 52.75 | 6.34 | 37.11 | 7.70 | 0.01 |
| **LDL IIIb** | 37.59 | 6.42 | 23.74 | 2.98 | <0.01 |
| **LDL IIIa** | 78.33 | 11.87 | 52.42 | 6.06 | <0.01 |
| **LDL IIb** | 75.58 | 10.66 | 51.93 | 10.86 | 0.01 |
| **LDL IIa** | 61.06 | 13.36 | 45.73 | 8.02 | 0.06 |
| **LDL I** | 72.15 | 21.50 | 82.62 | 16.52 | 0.41 |
| **IDL 2** | 54.95 | 6.45 | 62.92 | 13.09 | 0.02 |
| **IDL 1** | 54.95 | 6.45 | 62.92 | 13.09 | 0.25 |
| **VLDL sm** | 22.68 | 4.51 | 23.83 | 5.98 | 0.74 |
| **VLDL int** | 15.03 | 4.69 | 15.92 | 5.60 | 0.79 |
| **VLDL lg** | 3.96 | 1.90 | 4.49 | 2.52 | 0.72 |

**Supplementary Table 5:** LDL-APOA1 and LDL-APOA2 Fractional Clearance Rates on Placebo or rhLCAT Treatments

|  | Subject 1 | Subject 2 | Subject 3 | Subject 4 | Subject 5 | Mean | SD |
| --- | --- | --- | --- | --- | --- | --- | --- |
| **LDL-A1-FCR-Placebo** | 0.17 | 0.23 | 0.36 | 0.28 | 0.15 | 0.24 | 0.09 |
| **LDL-A1-FCR-rhLCAT** | 0.23 | 0.27 | 0.29 | 0.29 | 0.18 | 0.25 | 0.05 |
| **rhLCAT/Placebo diff** | 0.06 | 0.05 | -0.07 | 0.01 | 0.03 | 0.02 | 0.05 |
| **LDL-A2-FCR-Placebo** | 0.14 | 0.16 | 0.25 | 0.21 | 0.11 | 0.17 | 0.06 |
| **LDL-A2-FCR-rhLCAT** | 0.13 | 0.17 | 0.22 | 0.17 | 0.14 | 0.17 | 0.04 |
| **rhLCAT/Placebo diff** | -0.01 | 0.01 | -0.03 | -0.04 | 0.03 | -0.01 | 0.03 |

FCR: Fractional Clearance Rate calculated as pools/day, SD: standard deviation. Diff: Difference observed between rhLCAT and Placebo Treatment. Data for each study participant presented.

**Supplementary Table 6:** Sterol Levels During Placebo and rhLCAT Treatment. Levels of sterols in serum samples during Placebo and rhLCAT Periods. All levels were measured in the laboratory of Dr. Alice Lichtenstein^18^.

| **Sterols** | **Lathosterol**<br>**nmol/chol** | **Campesterol**<br>**nmol/chol** | **Beta-Sitosterol**<br>**nmol/chol** |
| --- | --- | --- | --- |
| **Placebo** |  |  |  |
| **1** | 30.1 | 363.3 | 2121.0 |
| **2** | 48.8 | 79.7 | 77.0 |
| **3** | 45.8 | 331.2 | 222.4 |
| **4** | 59.4 | 90.7 | 109.8 |
| **5** | 81.1 | 55.5 | 66.7 |
| **Mean** | **53.0** | **184.1** | **137.6** |
| SD | 18.9 | 149.9 | 74.5 |
| **Active** |  |  |  |
| **1** | 25.5 | 289.9 | 167.7 |
| **2** | 64.7 | 76.9 | 72.5 |
| **3** | 44.6 | 282.2 | 191.2 |
| **4** | 55.4 | 87.1 | 112.8 |
| **5** | 80.5 | 58.0 | 59.9 |
| **Mean** | **54.2** | **158.8** | **120.8** |
| SD | 20.7 | 116.6 | 57.6 |

**Supplementary Table 7:**
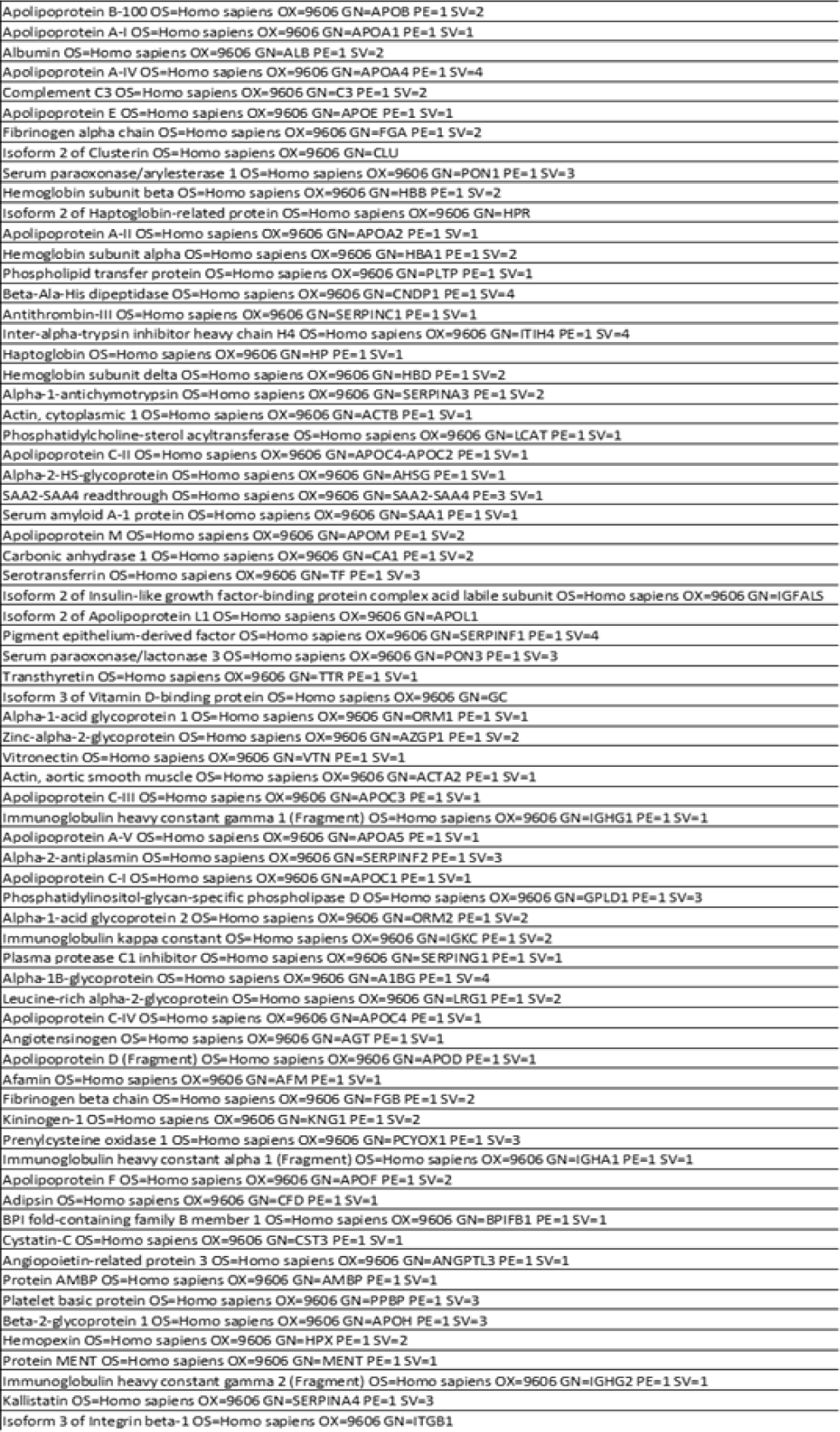
Identified HDL Associated Proteins

**Supplementary Table 8.**
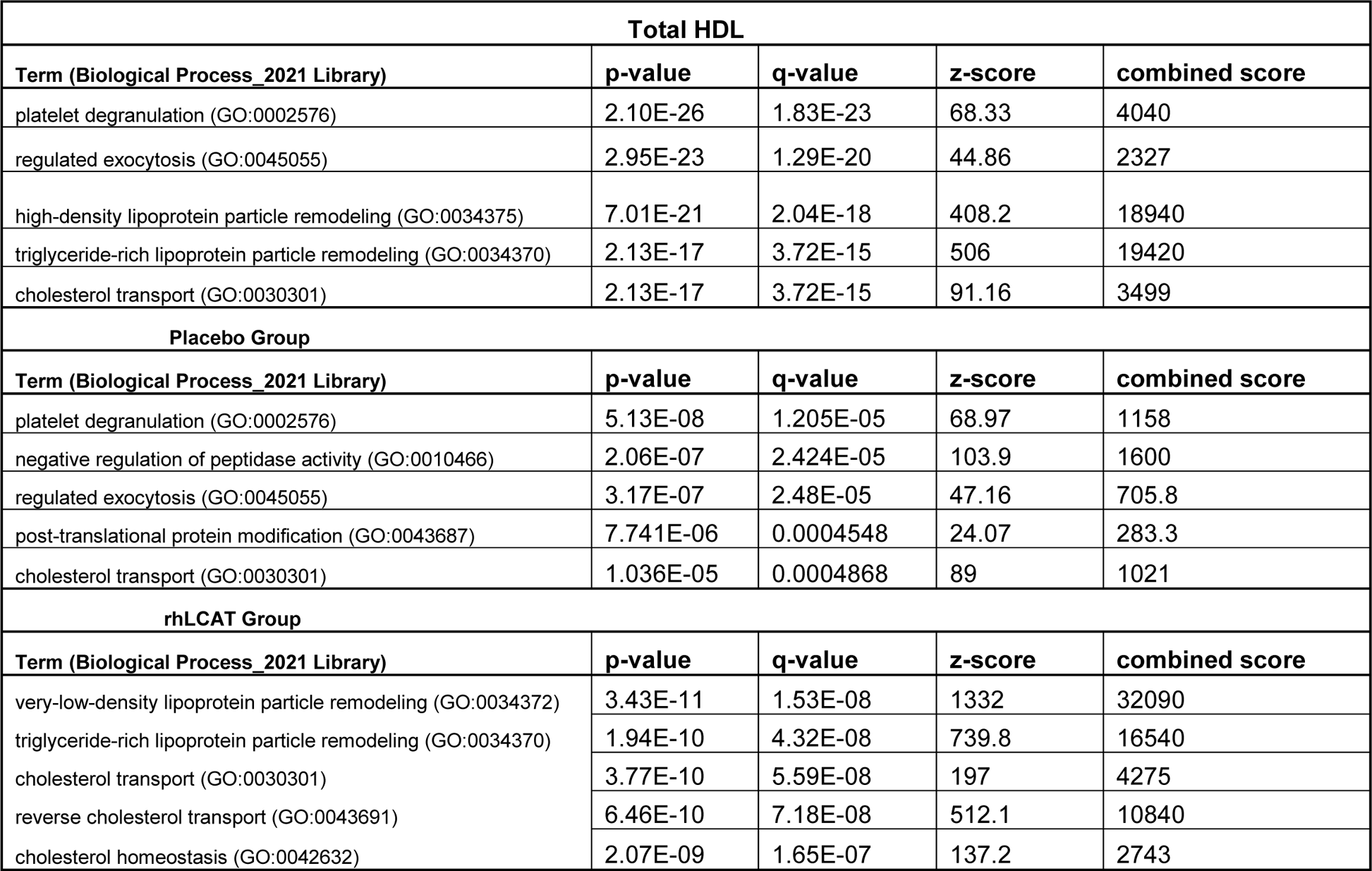
Top Biological Processes Identified by Gene Ontology Network Analysis. **Summary of the top** five biological processes for the three proteome queries (Total HDL, placebo group and rhLCAT group).

| **Total HDL** |  |  |  |  |
| --- | --- | --- | --- | --- |
| **Term (Biological Process_2021 Library)** | **p-value** | **q-value** | **z-score** | **combined score** |
| platelet degranulation (GO:0002576) | 2.10E-26 | 1.83E-23 | 68.33 | 4040 |
| regulated exocytosis (GO:0045055) | 2.95E-23 | 1.29E-20 | 44.86 | 2327 |
| high-density lipoprotein particle remodeling (GO:0034375) | 7.01E-21 | 2.04E-18 | 408.2 | 18940 |
| triglyceride-rich lipoprotein particle remodeling (GO:0034370) | 2.13E-17 | 3.72E-15 | 506 | 19420 |
| cholesterol transport (GO:0030301) | 2.13E-17 | 3.72E-15 | 91.16 | 3499 |
| **Placebo Group** |  |  |  |  |
| **Term (Biological Process_2021 Library)** | **p-value** | **q-value** | **z-score** | **combined score** |
| platelet degranulation (GO:0002576) | 5.13E-08 | 1.205E-05 | 68.97 | 1158 |
| negative regulation of peptidase activity (GO:0010466) | 2.06E-07 | 2.424E-05 | 103.9 | 1600 |
| regulated exocytosis (GO:0045055) | 3.17E-07 | 2.48E-05 | 47.16 | 705.8 |
| post-translational protein modification (GO:0043687) | 7.741E-06 | 0.0004548 | 24.07 | 283.3 |
| cholesterol transport (GO:0030301) | 1.036E-05 | 0.0004868 | 89 | 1021 |
| **rhLCAT Group** |  |  |  |  |
| **Term (Biological Process_2021 Library)** | **p-value** | **q-value** | **z-score** | **combined score** |
| very-low-density lipoprotein particle remodeling (GO:0034372) | 3.43E-11 | 1.53E-08 | 1332 | 32090 |
| triglyceride-rich lipoprotein particle remodeling (GO:0034370) | 1.94E-10 | 4.32E-08 | 739.8 | 16540 |
| cholesterol transport (GO:0030301) | 3.77E-10 | 5.59E-08 | 197 | 4275 |
| reverse cholesterol transport (GO:0043691) | 6.46E-10 | 7.18E-08 | 512.1 | 10840 |
| cholesterol homeostasis (GO:0042632) | 2.07E-09 | 1.65E-07 | 137.2 | 2743 |

**Supplementary Figure 1.**
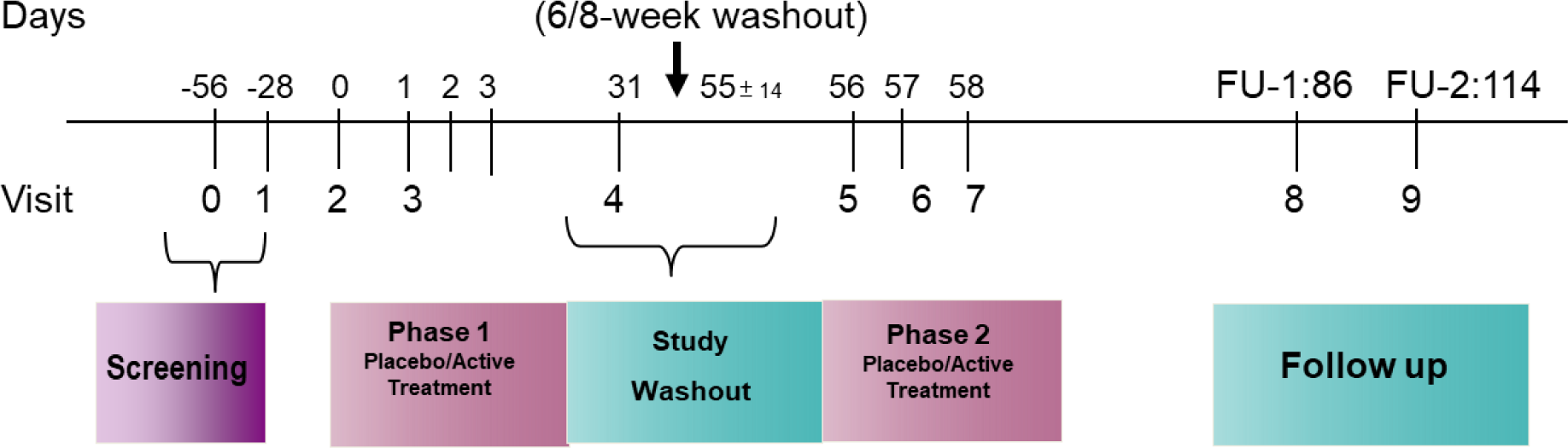
Study Design

**Supplementary Figure 2.**
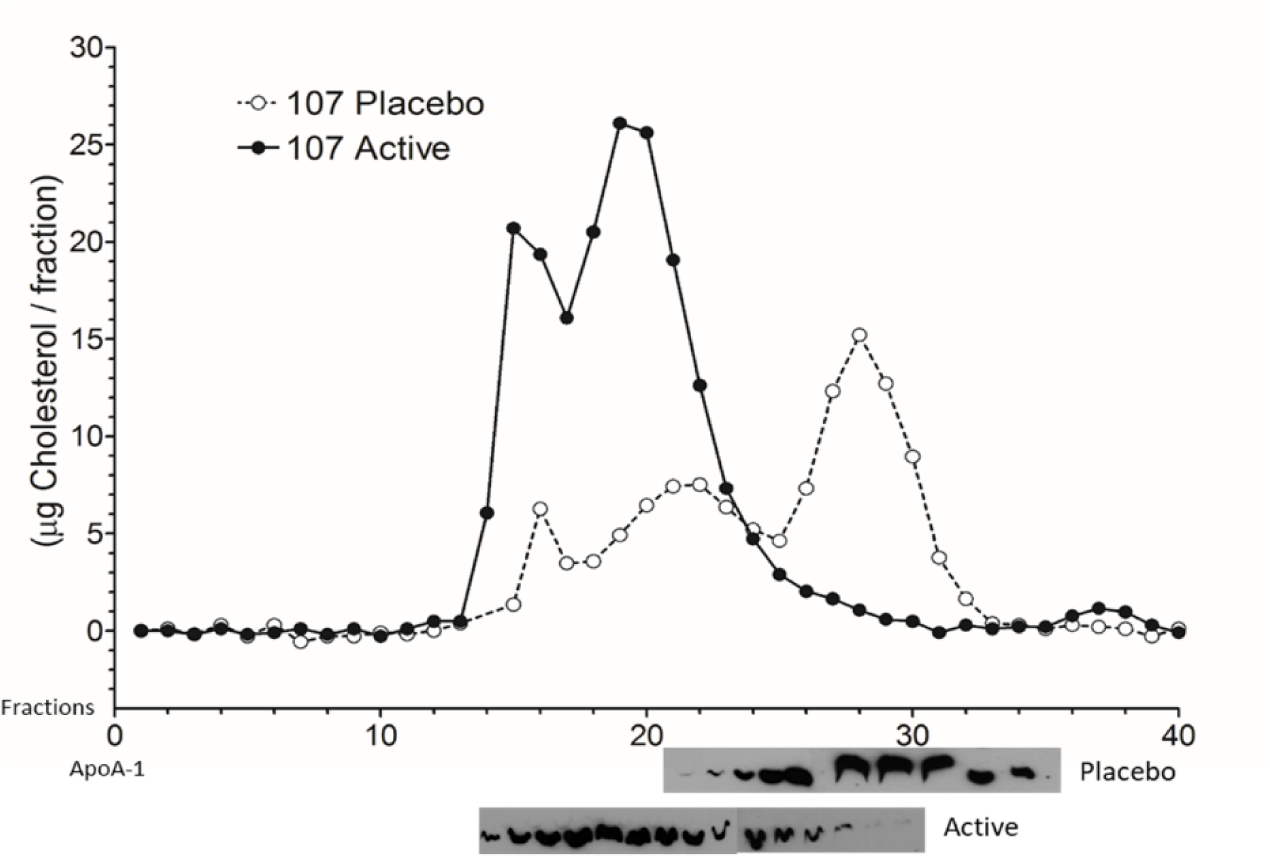
FPLC Size Distribution of Cholesterol and ApoA1 During Placebo and rhLCAT Treatment. Fast protein liquid chromatography (FPLC); active: rhLCAT: Apo: apolipoprotein. Plasma FPLC analyses demonstrating shift of APOA1 (lower western blot bands) and cholesterol from the HDL size-range (approx. fraction 25-35) into the LDL size-range (approx. fraction 15-25) during active treatment compared to placebo.

